# One-year subthalamic recordings in a patient with Parkinson’s disease under adaptive deep brain stimulation

**DOI:** 10.1101/2023.08.28.23294411

**Authors:** Laura Caffi, Luigi Michele Romito, Chiara Palmisano, Vanessa Aloia, Mattia Arlotti, Lorenzo Rossi, Sara Marceglia, Alberto Priori, Roberto Eleopra, Vincenzo Levi, Alberto Mazzoni, Ioannis Ugo Isaias

## Abstract

We present the clinical data and subthalamic recordings of a patient with Parkinson’s disease treated for one year with adaptive deep brain stimulation (aDBS). This novel stimulation mode, which adjusts the current amplitude linearly with respect to subthalamic beta power, produced a clinical benefit that was superior to the previous conventional stimulation that used constant, predefined parameters (cDBS). Compared with cDBS, the subthalamic beta amplitude was higher with aDBS and displayed larger daily fluctuations. Furthermore, subthalamic beta amplitude decreased during sleeping with respect to waking hours under aDBS. These data suggest a robust neuromodulatory mechanism of aDBS, with a clinical effect that was superior in this patient compared to cDBS. Our results open new perspectives for a restorative brain network effect of aDBS as a more physiologic, bidirectional, brain–computer interface.

## INTRODUCTION

Deep brain stimulation (DBS) of the subthalamic nucleus (STN) is a mainstay non-pharmacological treatment for selected Parkinson’s disease (PD) patients.^1,2^ Currently, the DBS paradigm is conventional DBS (cDBS), which is based on uninterrupted stimulation with clinically-determined fixed electrical settings (i.e., amplitude, pulse width, frequency, and wave-form), unrelated to the continuously changing functional state of the brain. Such cDBS programming aims to improve the main motor parkinsonian symptoms; however, the need to avoid unwanted, stimulation-related adverse effects, means that clinical responses may be suboptimal in some cases after cDBS .^3–6^ Adaptive DBS (aDBS) has the potential to optimize stimulation delivery through a responsive neuromodulation strategy, i.e., adapting stimulation parameters in a real-time manner by acquiring and elaborating symptom-specific and task-related biomarkers.^2,7^ To date, the most promising brain biomarkers in PD patients are the local field potentials (LFPs) recorded directly from implanted DBS electrodes. Strong oscillatory beta activity (13-30 Hz) of STN-LFPs could be a valid biomarker for bradykinesia and rigidity, as it is associated with the severity of PD-related motor symptoms^8^ and direct modulations on symptom improvement with levodopa administration^9^ and STN-DBS.^10^

Preliminary clinical evidence in short time windows suggests superior clinical efficacy of aDBS over cDBS in treating PD-related motor symptoms,^11–16^ with fewer stimulation side effects compared to cDBS.^17^ However, data on the long-term efficacy and safety of aDBS are still lacking, and its mechanism of action is still poorly understood. Accordingly, we describe the clinical and neurophysiological data collected over 11 months of follow-up in a patient with PD who underwent implantation of the AlphaDBS device (Newronika S.p.A.). Throughout this period, we evaluated the different effects of the two types of stimulation (aDBS or cDBS) on motor signs, sleeping or waking states, and dopaminergic medications, and tried to decode the specificities of the recorded subthalamic beta activity.

## RESULTS

### Consistent and sustained long-term clinical improvement with aDBS

Our patient is a male with an onset of parkinsonian signs (resting tremor and bradykinesia in the right hand) in his early 40s. Following the consistency of his clinical and symptomatologic evolution and the congruence of SPECT with FP-CIT imaging, he received a diagnosis of idiopathic PD and started therapy with dopamine agonists, levodopa, and iMAO agents, with excellent results. However, the development of severe motor fluctuations with peak-dose dyskinesias necessitated bilateral STN-DBS seven years after the onset of symptoms. Quadripolar electrodes (Medtronic 3389) were used, each connected to a Medtronic Activa SC 37603 implantable pulse generator (IPG), with remarkable clinical benefit. After about four years of treatment, the patient received the experimental AlphaDBS IPG as a replacement for the original battery-depleted Activa SC IPG (ClinicalTrials.gov identifier: NCT04681534; protocol NWK_AlphaDBS_FIM_2019, approved by the Fondazione IRCCS Istituto Neurologico Carlo Besta Local Ethics Committee). After implantation of the AlphaDBS IPG, the patient is currently being followed up for more than a year and a half.

A total of 168 days in aDBS+ and 47 days with cDBS+ were collected for our report (i.e., dopaminergic medication was continued, +). During this period, dopaminergic therapy was stable and maintained with levodopa/carbidopa 100/25 mg TID, opicapone 50 mg QD, and rasagiline 1 mg QD.

A switch from aDBS+ to cDBS+ was performed automatically by the device, following detection of a false positive sensing failure. Unaware of having returned to cDBS+, the patient asked for a visit because of a reduction in the clinical benefit of stimulation; upon checking, aDBS+ was reactivated. The aDBS+ settings were: C+1-, 2.6-3.9 mA, 130 Hz, 80 μs; C+8-, 2.6-3.0 mA, 130 Hz, 80 μs (**Figure 1**). The cDBS+ settings were: C+1-, 3.4 mA, 130 Hz, 80 μs; C+8-, 2.8 mA, 130 Hz, 80 μs. There was no statistical difference between the estimated total electrical energy delivered between DBS modalities.

**Figure 1.**
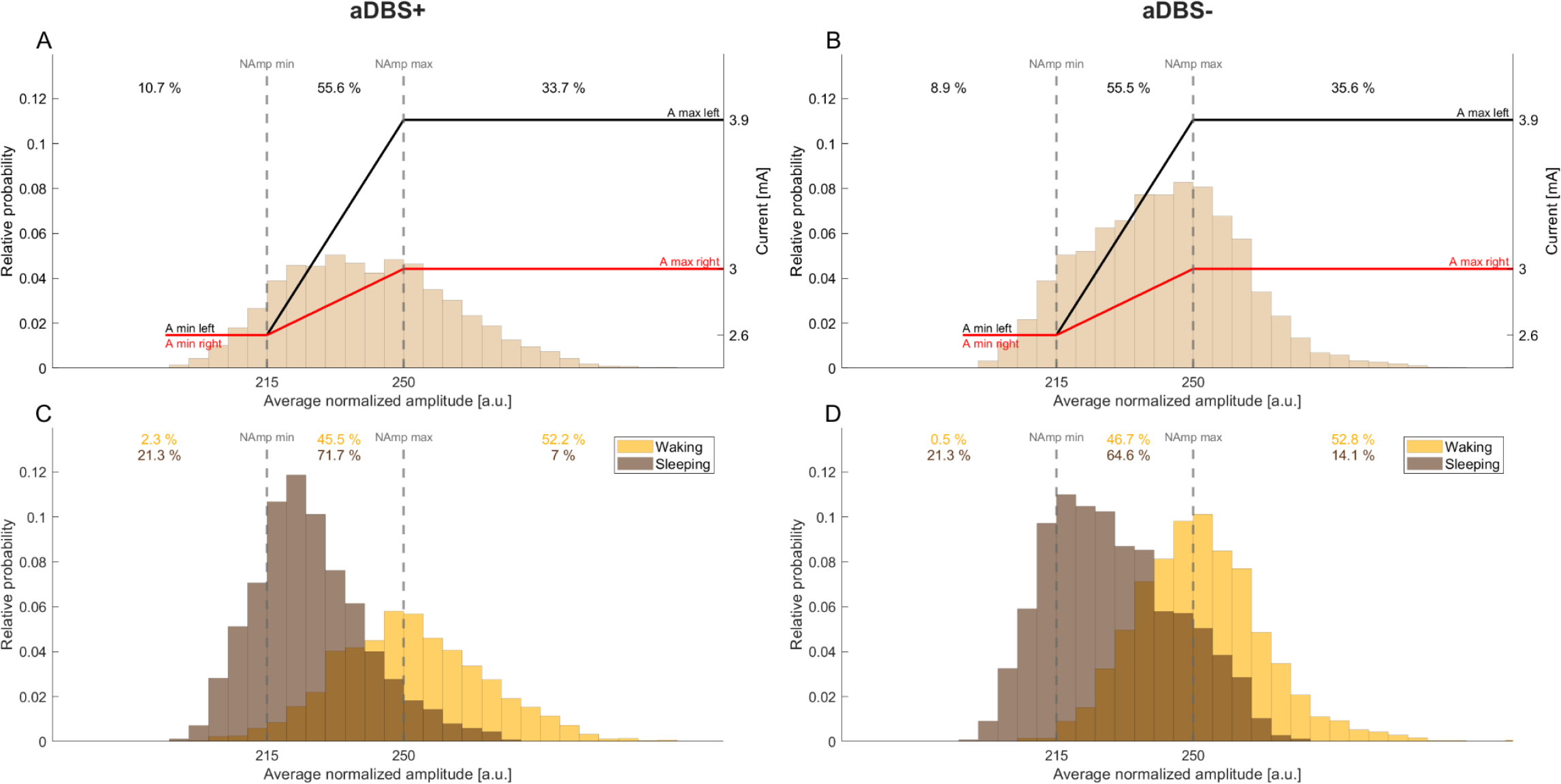
Principles of the AlphaDBS algorithm for current adjustment in adaptive mode. (A) Probability distribution (histogram) of the biomarker, which is used as the input signal for adjusting the current delivery, for the week in the aDBS+ condition that includes the representative day shown in **Figure 3C**. Specifically, the biomarker consists of an exponential moving average of the normalized amplitude samples (one per minute) recorded in the patient-specific beta frequency range (11-16 Hz) of the left STN (see STAR Methods). Vertical dotted lines represent the biomarker thresholds for current adjustment (NAmp min and NAmp max). Red and black solid lines represent the stimulation current at a specific reading. Specifically, if the average normalized amplitude at a given time is between NAmp min and NAmp max, the current is linearly adjusted within a predefined, clinically-effective range (Amin and Amax), independently for the two channels. Conversely, if the biomarker is below NAmp min, the current delivered remains Amin for the two hemispheres. Similarly, if the biomarker is above NAmp max, the current delivered remains Amax. Numbers on top show the time percentage of the average normalized amplitude being less than NAmp min, between NAmp min and NAmp max, and above NAmp max in the considered week. (B) Same as (A) for the aDBS-condition. (C) Probability distribution (histogram) of the biomarker for the same week displayed in (A) separately during waking (yellow) and sleeping (brown) in the aDBS+ condition. Vertical dotted lines represent the biomarker thresholds for current adjustment (NAmp min and NAmp max). Numbers on top show the time percentage of the average normalized amplitude being less than NAmp min, between NAmp min and NAmp max, and above NAmp max, in yellow and brown respectively for the waking and sleeping periods. (D) Same as (C) for the aDBS-condition. Abbreviations: a, adaptive; A, pre-defined, clinically-effective amplitude; c, conventional; DBS+, with dopaminergic medication; DBS-, without dopaminergic medication; NAmp, normalized beta amplitude; STN, subthalamic nucleus.

We also present data from a further 64-day period, during which time the patient requested to discontinue all dopaminergic medication (aDBS-condition), complaining of agitation and restlessness after taking them. Since then, the patient asked to maintain the discontinuation of drug therapy after judging the effect of aDBS-alone to be better than the aDBS+ condition.

System usability and technical issues caused sporadic data loss (45 days with unavailable data over the 11-month follow-up).

The patient consistently demonstrated a significant and stable benefit from bilateral STN-cDBS+. With this stimulation mode, the scores of the Unified Parkinson Disease Rating Scale (UPDRS) parts III and IV were 17/108 and 6/108 with both the Activa SC and the AlphaDBS devices. Additional improvement (UPDRS-III and -IV, respectively) was shown in aDBS+ by 18% and 50% and in aDBS-by 53% and 100% (absence of dyskinesias). During brief discontinuation (about 30 min) of DBS treatment, and after 12 hours of withdrawal of dopaminergic medications, the UPDRS-III score was 57/108.

The patient preferred the aDBS stimulation mode over cDBS for better control of PD-related motor symptoms; he also reported greater ease and enjoyment in all activities of daily living. He never complained of cognitive impairment, depression, anxiety or apathy, sleep problems, dysautonomia, hyposmia, or constipation. Genetic testing excluded common GBA, Park2, and PINK1 mutations.

### Long-term stable beta-band peak frequency

In the three stimulation conditions (cDBS+, aDBS+, and aDBS-), we acquired a unilateral (left) STN-LFPs spectrum every ten minutes (see STAR Methods). We analyzed separately the spectra recorded during waking (8am-10pm) and sleeping (midnight-6am) (see STAR Methods). We identified three spectral peaks in the 7-34 Hz range, which remained stable across the whole recording periods and stimulation conditions (values reported in Hz as median [first quartile, third quartile]: first peak: 8.2 [7.8, 8.3] (waking) and 8.2 [7.7, 8.4] (sleeping); second peak: 12.7 [12.5, 12.9] (waking) and 12.3 [12.1, 12.5] (sleeping); third peak: 24.7 [23.9, 25.7] (waking) and 24.8 [24.3, 25.5] (sleeping) (**Figure 2**).

**Figure 2.**
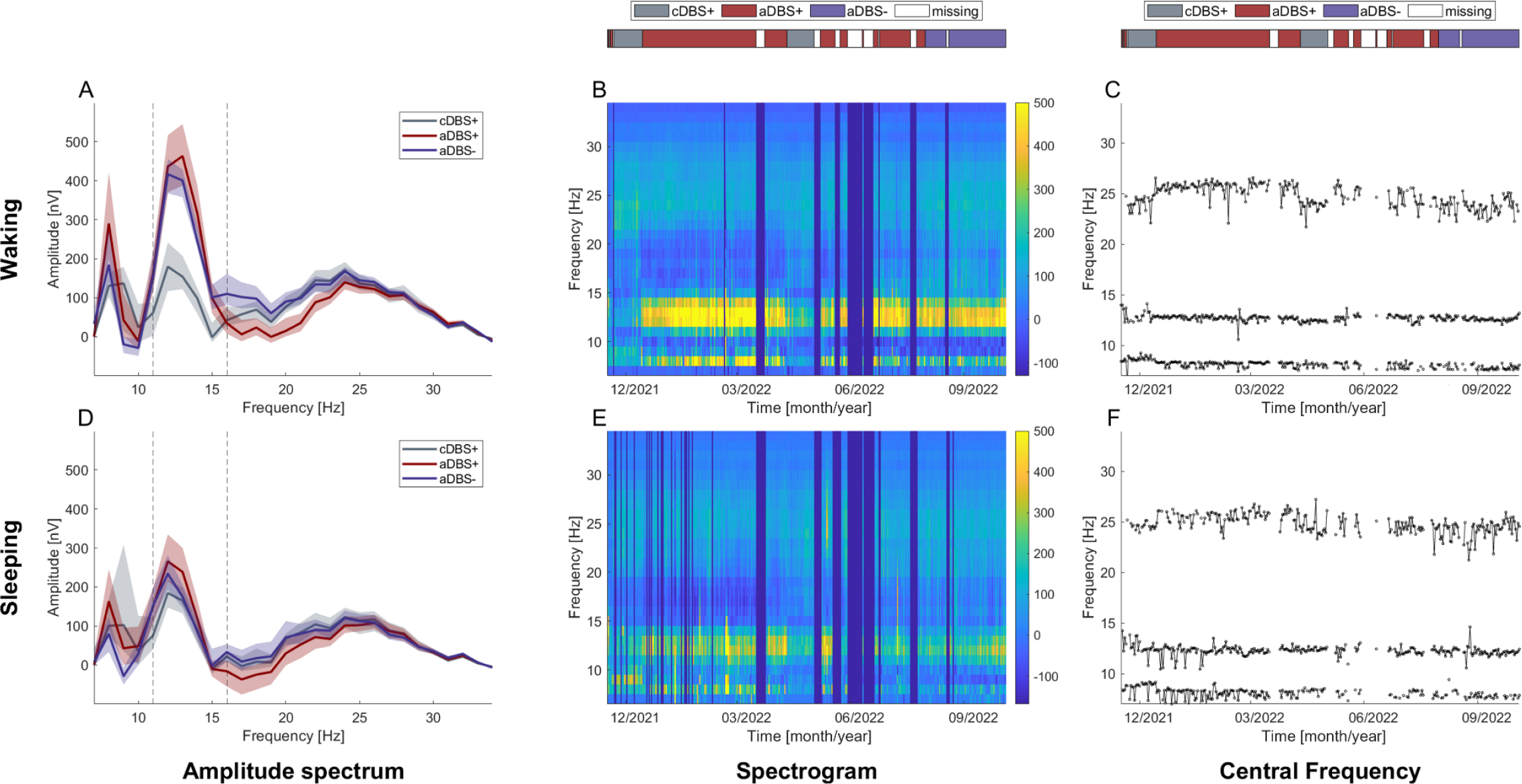
Recording of the STN-LFP amplitude spectrum over 11 months. (A) Median (solid line) of the daily mean amplitude spectra throughout the 11 months of recording in the three treatment conditions (cDBS+ in grey, aDBS+ in dark red, and aDBS-in purple) during waking. The daily mean spectra are cleaned from 1/*f*^*n*^ noise (see STAR Methods). The dashed area is bounded by the first and third quartiles of the daily mean amplitude spectra. (B) Spectrogram of daily mean amplitude spectra cleaned from 1/*f*^*n*^ noise (see STAR Methods) during waking. Blue vertical lines correspond to missing or removed data periods due to residual spectral artifacts after neural power law component removal (see STAR Methods). (C) Time course of the central frequency of the three Gaussian peaks identified in each daily mean amplitude spectrum (see STAR Methods) during waking. (D) Same as (A) during sleeping. (E) Same as (B) during sleeping. (F) Same as (C) during sleeping. Abbreviations; a, adaptive; c, conventional; DBS+, with dopaminergic medication; DBS-, without dopaminergic medication; LFPs, local field potentials; STN, subthalamic nucleus.

### Sleep-wake variation in beta amplitude was larger in aDBS than in cDBS

We computed the daily median and interquartile range of the patient-specific STN-LFPs beta frequency range amplitude (BFRA) from the spectra acquired every ten minutes, calculated separately for the waking and sleeping periods (see STAR Methods).

We observed a statistically significant interaction between treatment (cDBS+, aDBS+, and aDBS-) and activity level (waking and sleeping) in determining the daily median BFRA (mixed ANOVA on the ranks, see STAR Methods: F(2,296)=21.23, p<0.001). Simple main effects analysis showed that treatment (p<0.001) and activity level (p<0.001) both had a statistically significant effect on the daily median BFRA. During waking, this value was significantly higher in aDBS+ and aDBS-than in cDBS+ (Kruskal-Wallis: p<0.001, **Figure 3 and Table 1**). During sleeping, the daily median BFRA did not differ between treatments (Kruskal-Wallis: p=0.15). A statistically significant difference in the daily median BFRA between waking and sleeping was observed for both the aDBS+ and aDBS-conditions (Wilcoxon signed-rank: p<0.001) but not for cDBS+ (**Figure 3 and Table 1**).

**Figure 3.**
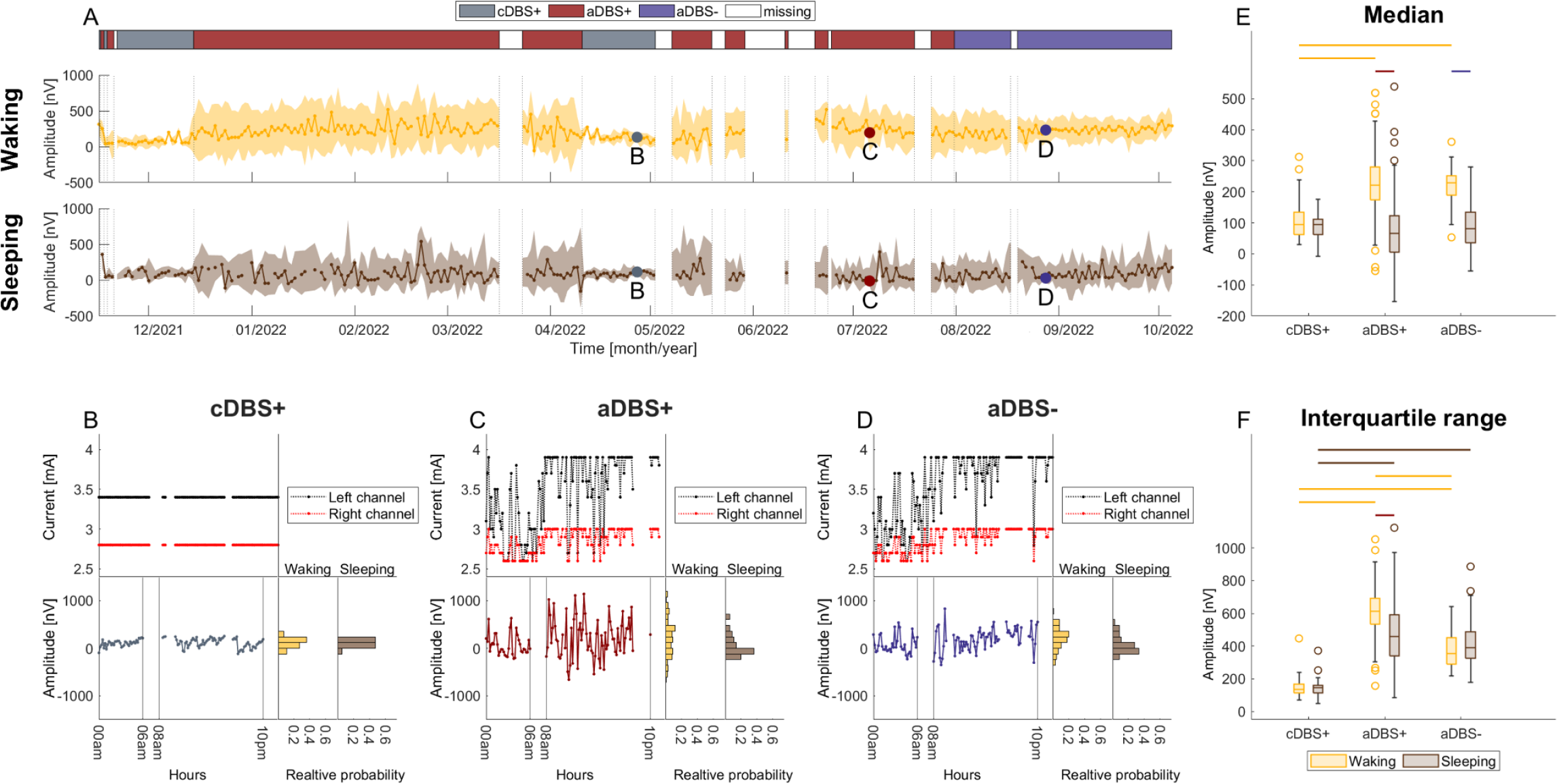
Evolution of STN-LFP amplitude in the patient-specific beta range during 11 months of recording (A) Total evolution of the daily median BFRA during waking and sleeping (solid line). The shadowed area is bound by the daily first and third quartile of the BFRA. Vertical dotted lines represent the time points in which the treatment condition changed, as displayed in the legend. Grey, dark red, and purple dots mark the representative days shown respectively in (B), (C), and (D). (B) Top left: daily evolution of the stimulation current for the left (black) and right (red) channel for a representative day in cDBS+. Bottom left: daily evolution of the BFRA for the same day in cDBS+. Waking (8am-10pm) and sleeping (midnight-6am) periods are separated by vertical solid lines. Bottom right: probability distribution (histogram) representing the distribution of the BFRA sampled every ten minutes for the same representative day in cDBS+ separately for the waking (yellow) and sleeping (brown) periods. (C) Same as (B) for a representative day in aDBS+. (D) Same as (B) for a representative day in aDBS-. (E) Boxplot of the daily median BFRA during waking (yellow) and sleeping (brown) with cDBS+, aDBS+, and aDBS-(values reported in nV as median [first quartile, third quartile]). Waking: cDBS+: 94.6 [61.4, 134.3], aDBS+: 220.3 [174, 279.5], aDBS-: 228.5 [189, 252.2]. Sleeping: cDBS+: 94.5 [62, 110.6], aDBS+: 65.6 [5.6, 121.9], aDBS-: 80.7 [36.6, 135]). The significance level was set to 0.05. Top horizontal lines define significant differences (dashed line: 0.001<p<0.01; solid line: p<0.001). (F) Same as (E) for the interquartile range of the BFRA. Waking: cDBS+: 133.8 [114.9, 167.1], aDBS+: 612 [533.1, 690.3], aDBS-: 354 [287.8, 452.2]. Sleeping: cDBS+: 144.6 [112, 161.3], aDBS+: 459.3 [339.8, 593.1], aDBS-: 390.5 [325.3, 488.4]). Abbreviations: a, adaptive; BFRA, beta frequency range amplitude; c, conventional; DBS+, with dopaminergic medication; DBS-, without dopaminergic medication; LFPs, local field potentials; STN, subthalamic nucleus.

**Table 1.**
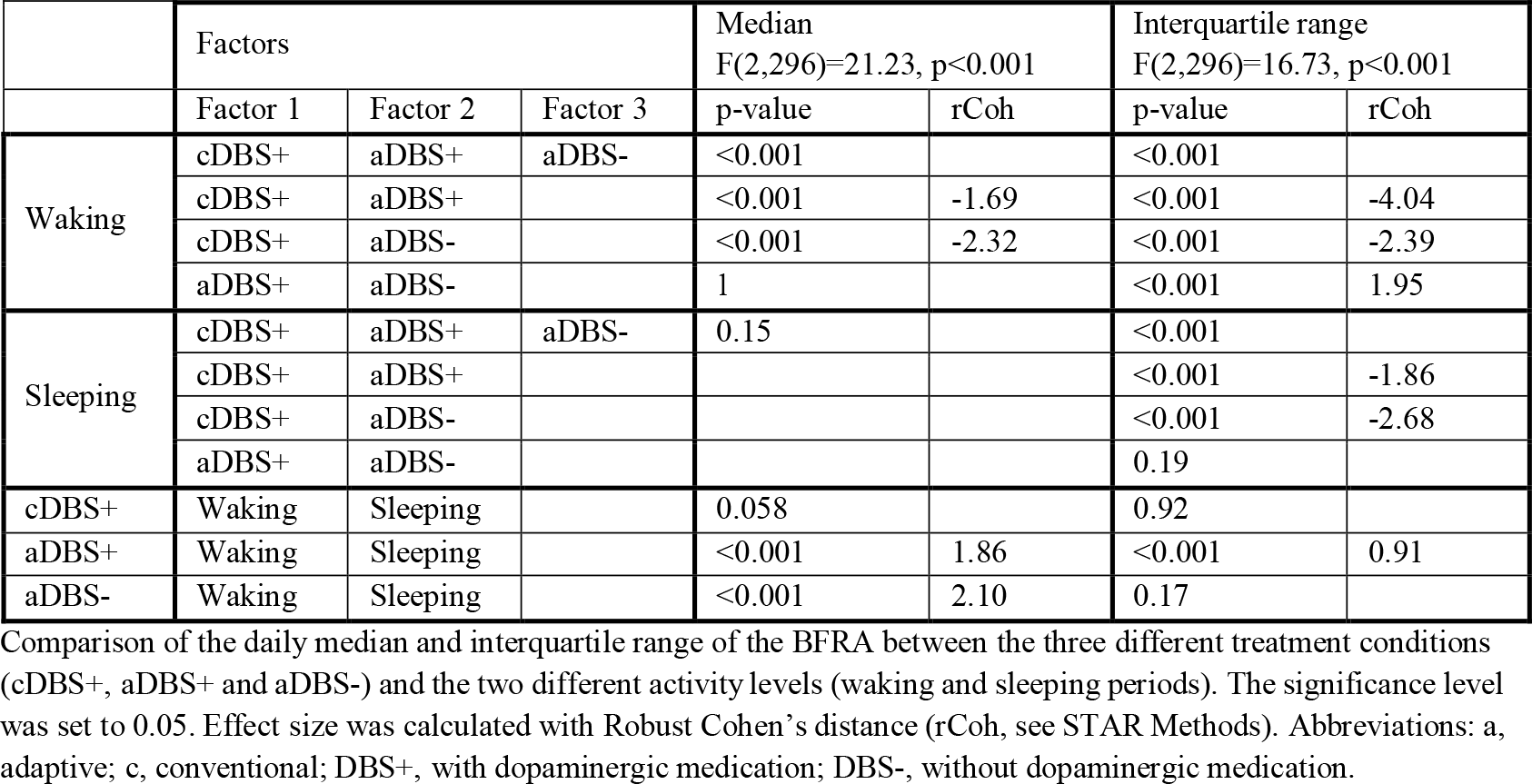
Statistical comparisons in the patient-specific beta range.

### STN beta amplitude variability was larger in aDBS than in cDBS

We found a significant interaction between the treatment and activity level in determining the daily interquartile range of BFRA (mixed ANOVA on the ranks: F(2,296)=16.73, p<0.001, **Figure 3**). Both factors had a statistically significant effect (p<0.001). During waking, the interquartile range was significantly higher in aDBS+ than in cDBS+ and aDBS-, and in aDBS-compared to cDBS+ (Kruskal-Wallis: p<0.001, **Figure 3 and Table 1**). The same ranking was replicated during sleeping, but with no significant difference between aDBS+ and aDBS-(Kruskal-Wallis: p<0.001, **Figure 3 and Table 1**). For the daily BFRA interquartile range, we also observed a reduction in sleeping compared to waking but in aDBS+ only (Wilcoxon signed-rank: p<0.001, **Figure 3 and Table 1**). In both aDBS+ and aDBS-, the median BFRA contained significant information about the waking and sleeping condition (0.32 and 0.42 bits respectively, bootstrap test, p<0.05, see Star Methods and **Figure S1**), but its interquartile range only carried significant information in aDBS+ (0.20 bits, p<0.05 vs 0.02 bits p>0.1 in aDBS-, **Figure S1**). Of relevance, neither feature carried information about the waking and sleeping condition in cDBS+ (0.01 and 0.01, p>0.1, **Figure S1**).

Of note, all analyses were performed on the patient-specific beta frequency range (11-16 Hz, see STAR Methods). By repeating the same analyses for the conventional beta frequency bands, very similar results were obtained in the low beta band (13-20 Hz, **Figure S2 and Table S1**), while in the high beta band (21-30 Hz) a similar pattern was only seen for the daily interquartile range of the STN-LFP amplitude during waking (**Figure S3 and Table S2**).

## DISCUSSION

The aDBS paradigm described, with linear current modulation, proved superior in the long-term control of motor symptoms and improvement of patient wellbeing compared with cDBS. From a pathophysiological point of view, our data showed overall stability of the STN beta-frequency peaks over time. Of relevance, we showed a difference in modulation of the beta amplitude and fluctuations related to waking and sleeping with aDBS compared with cDBS. This applies to the patient-specific frequency range and to conventional frequency bands, in particular the low beta band.

The subjective and objective superior clinical benefit of aDBS over cDBS, and the gradual discontinuation of all pharmacological treatment, should be considered as a novel condition – possibly a functional recovery – permitted by a putative (re)activation of compensatory basal ganglia-thalamic-cortical circuitries yet to be identified. A brain–computer aDBS interface, driven by linear algorithms for stimulation delivery and adaptation, could theoretically determine not only an immediate benefit on the parkinsonian symptoms, determined by the detection of specific changes in symptom-related biomarkers, but also a more significant long-term comprehensive clinical improvement, possibly permitted by the restoration of more physiological key neural activities (e.g., beta oscillations). This perspective is reflected in our observations, which show greater beta oscillatory activity in aDBS than in cDBS, paralleling greater clinical benefit over time.

While cDBS would exclusively retain a suppressive action on pathological neural activity, aDBS may permit and/or promote a more profound integration of the functional and informative^18,19^ components of the beta oscillations, and a more natural circadian rhythm of these oscillations, while retaining the positive effect on parkinsonian symptoms. In line with this hypothesis, we think that the reduced subthalamic beta activity during sleep depends on the lower involvement of the STN compared with voluntary motor control in the nocturnal regulation of sleep-related rhythmic and homeostatic processes. This reasoning may also justify the different responses to chronic (over months) or acute aDBS (during a pharmacological test with levodopa).^11,20^ In the latter case, only the suppressive response on akinetic-rigid signs would be evident.

Our recordings also demonstrate the overall stability of beta-band peaks over time. This result is important for current aDBS algorithms because it testifies to the capability of correctly monitoring power modulations over an established range of frequencies. However, we must recognize that we were unable to correlate subthalamic activity and kinematic parameters to allow description of specific changes in neural activity (e.g., frequency modulation^19^) under certain conditions, such as walking. Indeed, there is increasing evidence that the beta peak frequency is an important and functionally-relevant parameter of oscillatory activity both at a cortical^21^ and subcortical^19^ level.

We conclude that our clinical case provides preliminary first evidence of the clinical efficacy of aDBS over one year, paving the way for new studies with more reliable neuromodulation strategies based on continuous bidirectional brain– computer communication – not only to better tailor symptomatic improvement, but also to possibly (re)activate new compensatory brain resources.

## Data Availability

All data produced in the present study are available upon reasonable request to the authors

## ACKNOWLEDGMENTS

The study was funded by the European Union - Next Generation EU - NRRP M6C2 - Investment 2.1 Enhancement and strengthening of biomedical research in the NHS, and by the Fondazione Grigioni per il Morbo di Parkinson. IUI and CP were supported by the Deutsche Forschungsgemeinschaft (DFG, German Research Foundation) Project-ID 424778381 - TRR 295. IUI was supported by a grant from the New York University School of Medicine and The Marlene and Paolo Fresco Institute for Parkinson’s and Movement Disorders, which was made possible with support from Marlene and Paolo Fresco.

We are grateful to many colleagues for their help in this line of research. In particular, we would like to thank: Salvatore Bonvegna, Elena Contaldi, and Manuela Pilleri of the Parkinson Institute Milan, ASST G. Pini-CTO; Nicoló Pozzi and Ibrahem Hanafi of the University Hospital Würzburg; Sara Rinaldo and Nico Golfrè Andreasi of the Parkinson and Movement Disorders Unit, Fondazione IRCCS Istituto Neurologico Carlo Besta, Milan; Costanza Conti of Newronika S.p.A..

## AUTHOR CONTRIBUTIONS

LC, LuR, CP, MA, LR, AM, IUI: Conceptualization; LC, LuR, CP, VA, VL, IUI: Data collection; LC, MA, CP: Data curation; LC, CP, AM, IUI: Formal analysis; MA, LuR, RE, AP, IUI: Funding acquisition; LC, LuR, CP, MA, LR, AM, IUI: Methodology; LR, RE, SM, AP, AM, IUI: Resources; CP, MA, AM, IUI: Supervision; LC, LuR, AM, IUI: Writing - original draft; CP, VA, MA, LR, SM, AP, VL, RE: Writing - review & editing.

## DECLARATION OF INTERESTS

VA, MA, and LR are employees and shareholders of Newronika S.p.A. IUI is a Newronika S.p.A. consultant and shareholder. SM and AP are founders and shareholders of Newronika S.p.A. IUI and RE received funding for research activities from Newronika S.p.A. IUI is Adjunct Professor at the Department of Neurology, NYU Grossman School of Medicine.

We support inclusive, diverse, and equitable conduct of research.

## STAR METHODS

### Resource availability

#### Lead contact

Further information and requests for resources should be directed to and will be fulfilled by the lead contact, Ioannis U. Isaias.

#### Material availability

- The AlphaDBS implantable pulse generator (Newronika S.p.A.) is commercially available for cDBS and is an experimental device for aDBS.

#### Data and code availability

- LFPs recorded with the AlphaDBS device cannot be deposited in a public repository because they can be traceable to the identity of the subject. They will be made available upon reasonable request to the lead contact.
- No original method has been developed. All analyses were performed in MATLAB 2021a and MATLAB 2023a (The MathWorks Inc., Natik, Massachusetts, USA) with standard functions.
- Any additional information required to reanalyze the data reported in this paper is available from the lead contact upon request.

### Methods details

#### Adaptive paradigm of the AlphaDBS device

The AlphaDBS device in aDBS mode^22^ applies a linear algorithm that provides a stimulation amplitude (within a predefined, clinically-effective range) based on an exponential moving average with a time constant of 50 s of the amplitude samples (one per second), recorded in a patient-specific, beta frequency range in one STN. This is performed continuously, with one sample (1 s recording) entering and exiting the average calculation. The stimulation frequency and pulse width remain fixed. The left STN and the recording contact pair 0-2 were chosen in our patient, as it showed the most prominent beta peak among all contact pairs. The frequency range monitored (11-16 Hz) was defined as ±2.5 Hz centered to the highest beta peak. Beta amplitude samples were normalized over the total power in the 5-34 Hz range.

The normalized beta amplitude distribution in this frequency range was initially monitored for three days with cDBS+ and checked at different follow-up visits. This allowed identification of the normalized beta amplitude limits (NAmp min and NAmp max) by which the stimulation current was to be delivered (**Figure 1**).

The two stimulation current thresholds were clinically defined as the amplitude (Amin) providing 40-50% clinical benefit in meds-off state (i.e., titrating up the stimulation current in the morning after overnight suspension of all dopaminergic drugs) and the maximum amplitude (Amax) in the absence of side effects in the meds-on condition (i.e., titrating up the stimulation current at 60 min after 100+25 mg levodopa+carbidopa intake) (**Figure 1**).

#### Spectral analysis

With active stimulation, the AlphaDBS device saved the stimulation current and the average subthalamic amplitude spectrum (from 5-34 Hz with 1 Hz resolution) every ten minutes.^22^

We analyzed the waking and sleeping periods separately, since a reduction of subthalamic beta power during sleep has been reported in patients treated with cDBS.^23^ The same time windows for waking (8am-10pm) and sleeping (midnight-6am), chosen according to the patient’s daily routine, were used for each day. Recordings from 6am to 8am and from 10pm to midnight were excluded because of the differences in the patient’s daily schedule (e.g., time of falling asleep, etc.).

To investigate relevant spectral peaks, we cleaned the amplitude spectra from 1/*f*^*n*^ noise as follows. For each day, we computed two average spectra, one for waking and one for sleeping, by averaging the amplitude spectra acquired from 8am to 10pm and from midnight to 6am, respectively. The daily average spectra showed an aperiodic component superimposed to the oscillatory peaks. Consequently, after identification of the aperiodic component starting frequency (7 Hz) by visual inspection, the waking/sleeping average spectra for each day was decomposed in the two components, aperiodic and periodic; these were modeled respectively as exponential functions in semi-logarithmic amplitude-space with characteristic offset, slope, and bend, and Gaussian functions with characteristic central frequency, amplitude, and width.^24^ The quality of the decomposition was visually inspected and days presenting residual spectral artifacts after subtraction of the aperiodic component were removed by the analysis. Moreover, the presence and stability of the Gaussian peaks were inspected across days and conditions. For further analysis, for each day the daily waking or sleeping periodic components were subtracted from each ten-minute amplitude spectrum acquired during the day.

#### Quantification and statistical analysis

Within each day, separately for the waking and sleeping periods, we calculated the median and interquartile range of the BFRA of each ten-minute amplitude spectrum acquired during the day. The effect of treatment (cDBS+, aDBS+, and aDBS-) and activity level (waking and sleeping) was evaluated using mixed ANOVA on the ranked data.^25^

Subsequently, a single main effect analysis was conducted with non-parametric tests after checking for normality with the Anderson-Darling test. Differences between the treatment conditions, separately for waking and sleeping periods, were assessed using the Kruskal-Wallis test. Post-hoc analysis was conducted with Tukey’s honestly significant difference procedure. The effect of the activity level (waking and sleeping) in the different treatment conditions was conducted through Wilcoxon signed-rank test. The statistical significance was set at 0.05. The effect size was calculated through Robust Cohen’s distance (rCoh) obtained from the Cohen’s distance by replacing population means with 20% trimmed means and the population standard deviation with the square root of a 20% Winsorized variance.^26^ Mutual information between the waking/sleeping condition and BFRA features was computed based on standard methods using Panzeri-Treves method for bias correction and bootstrap test for significance.^27^

