## Supplemental Material for "One-year subthalamic recordings in a patient with Parkinson’s disease under adaptive deep brain stimulation"

### SUPPLEMENTAL MATERIALS

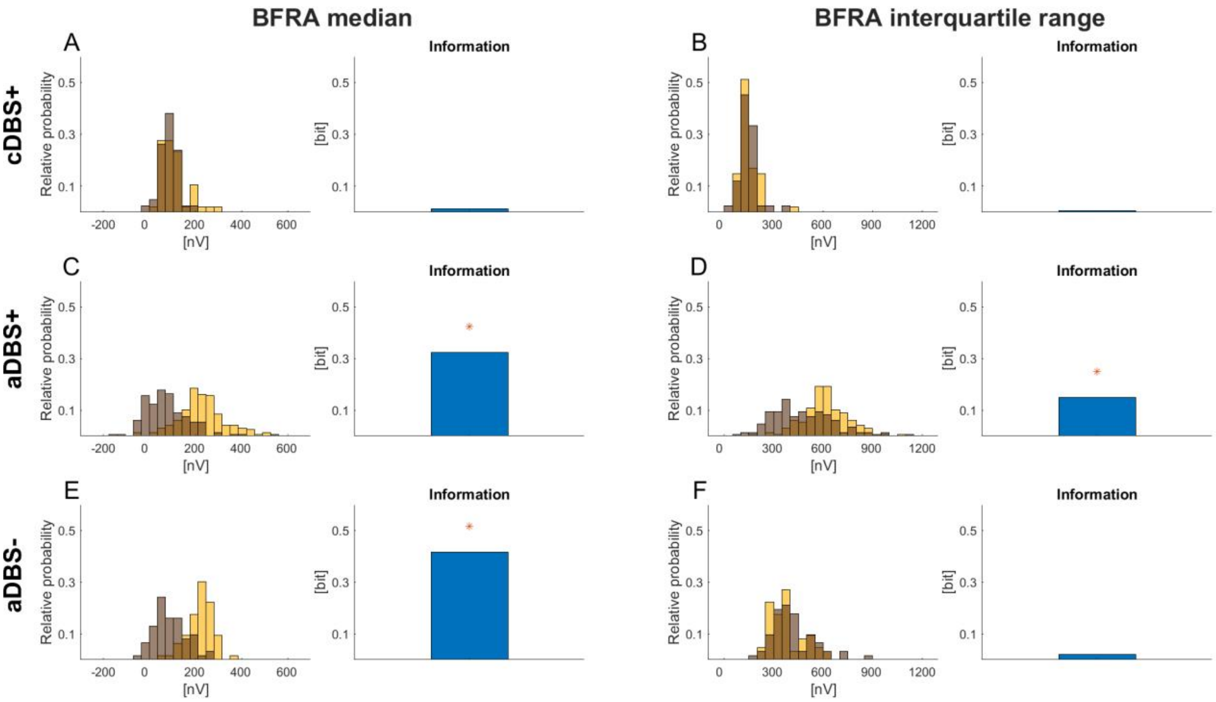


**Figure S1. Information carried by the BFRA median and interquartile ranges on the waking/sleeping condition.**

(A) Left: distribution of the median BFRA for all days in the cDBS+ condition. Right: information carried by the median BFRA on the waking/sleeping condition (see STAR Methods).
(B) Same as (A) for the interquartile range of the BFRA in cDBS+.
(C) Same as (A) for aDBS+ condition.
(D) Same as (B) for aDBS+ condition.
(E) Same as (A) for aDBS- condition.
(F) Same as (B) for aDBS- condition.
Abbreviations: a, adaptive; BFRA, beta frequency range amplitude; c, conventional; DBS+, with dopaminergic medication; DBS-, without dopaminergic medication.

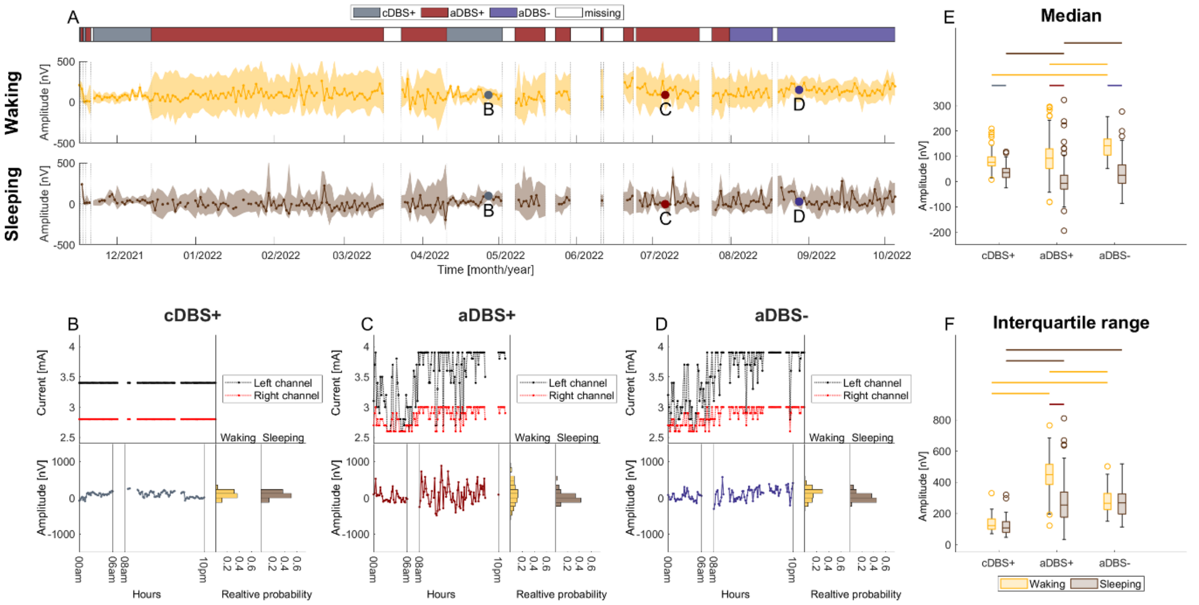


**Figure S2. Evolution of STN-LFP amplitude in the low beta band (13-20 Hz) during 11 months of recording. As Figure 2 of the main text, but for the low-beta band.**

(A) Total evolution of the daily median low beta amplitude during waking and sleeping (solid line). The shadowed area is bound by the daily first and third quartile of the low beta amplitude. Vertical dotted lines represent the time points in which the treatment condition changed, as displayed in the top panel. Grey, dark red, and purple dots mark the representative days shown respectively in (B), (C), and (D).
(B) Top left: daily evolution of the stimulation current for the left (black) and right (red) channel for a representative day in cDBS+. Bottom left: daily evolution of the low beta amplitude for the same representative day in cDBS+. Waking (8am-10pm) and sleeping (midnight-6am) periods are separated by vertical solid lines. Bottom right: probability distribution (histogram) representing the distribution of the low beta amplitude for the same representative day in cDBS+ separately for the waking (yellow) and sleeping (brown) periods.
(C) Same as (B) for a representative day in aDBS+.
(D) Same as (B) for a representative day in aDBS-.
(D) Boxplot of the daily median low beta amplitude during waking (yellow) and sleeping (brown) in cDBS+, aDBS+, and aDBS- (values reported in nV as median [first quartile, third quartile]). Waking: cDBS+: 77 [61.5, 97], aDBS+: 92.7 [51.8, 129.5], aDBS-: 141.3 [104.5, 168.3]. Sleeping: cDBS+: 35.8 [16, 51.2], aDBS+: -6 [-29.1, 25.6], aDBS-: 24.4 [-6.9, 65.3]). The significance level was set to 0.05. Top horizontal lines define significant differences (dashed line: $0.001<p<0.01$; solid line: $p<0.001$).
(E) Same as (C) for the interquartile range of the low beta amplitude. Waking: cDBS+: 122.1 [99.5, 165.5], aDBS+: 449.8 [386.5, 514.5], aDBS-: 265 [225, 326.7]. Sleeping: cDBS+: 107.2 [78, 146.8], aDBS+: 253 [174.9, 336.5], aDBS-: 269.1 [194.7, 324.6]).
Abbreviations: a, adaptive; c, conventional; DBS+, with dopaminergic medication; DBS-, without dopaminergic medication; LFPs, local field potentials; STN, subthalamic nucleus.

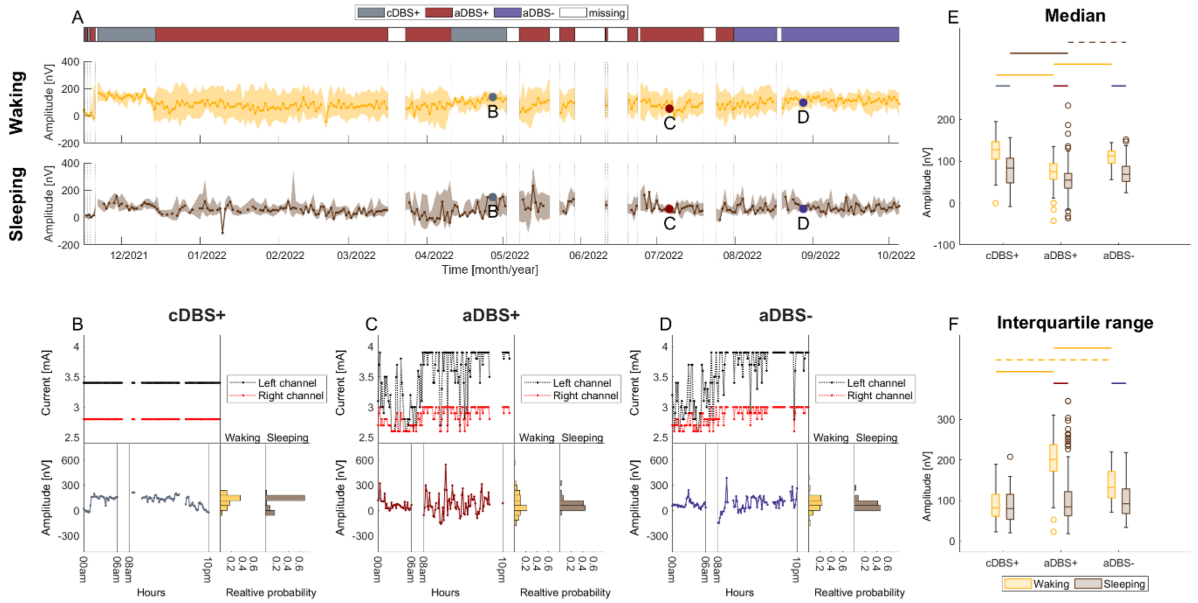


**Figure S3. Evolution of the STN-LFP amplitude in the high beta band (21-30 Hz) during 11 months of recording. As Figure 2 of the main text, but for the high beta band.**

(A) Total evolution of the daily median high beta amplitude during waking and sleeping (solid line). The shadowed area is bound by the daily first and third quartile of the high beta amplitude. Vertical dotted lines represent the time points in which the treatment condition changed, as displayed in the top panel. Grey, dark red, and purple dots mark the representative days shown respectively in (B), (C), and (D).
(B) Top left: daily evolution of the stimulation current for the left (black) and right (red) channel for a representative day in cDBS+. Bottom left: daily evolution of the high beta amplitude for the same representative day in cDBS+. Waking (8am-10pm) and sleeping (midnight-6am) periods are separated by vertical solid lines. Bottom right: probability distribution (histogram) representing the distribution of the high beta amplitude for the same representative day in cDBS+ separately for the waking (yellow) and sleeping (brown) periods.
(C) Same as (B) for a representative day in aDBS+.
(D) Same as (B) for a representative day in aDBS-.
(D) Boxplot of the daily median high beta amplitude during waking (yellow) and sleeping (brown) in cDBS+, aDBS+, and aDBS- (values reported in nV as median [first quartile, third quartile]). Waking: cDBS+: 127.7 [104.5, 146.9], aDBS+: 75.4 [56.6, 93.8], aDBS-: 112.9 [95.4, 124.2]. Sleeping: cDBS+: 84 [48.2, 107.2], aDBS+: 54.9 [35.9, 71.1], aDBS-: 68.3 [51.2, 88]). The significance level was set to 0.05. Top horizontal lines define significant differences (dashed line: $0.001<p<0.01$; solid line: $p<0.001$).
(D) Same as (C) for the interquartile range of the high beta amplitude. Waking: cDBS+: 82.1 [61.4, 115], aDBS+: 200.7 [172.2, 237.5], aDBS-: 131.5 [106.5, 171.3]. Sleeping: cDBS+: 79.7 [53, 114.7], aDBS+: 84.6 [61.8, 122.1], aDBS-: 91.9 [68, 128.5]).
Abbreviations: a, adaptive; c, conventional; DBS+, with dopaminergic medication; DBS-, without dopaminergic medication; LFPs, local field potentials; STN, subthalamic nucleus.

| | **Factors** | | | **Median** F(2,296)=12.12, p<0.001 | | **Interquartile range** F(2,296)=28.18, p<0.001 | |
|---|---|---|---|---|---|---|---|
| | **Factor 1** | **Factor 2** | **Factor 3** | **p-value** | **rCoh** | **p-value** | **rCoh** |
| Waking | cDBS+ | aDBS+ | aDBS- | <0.001 | | <0.001 | |
| | cDBS+ | aDBS+ | | 0.24 | | <0.001 | -3.66 |
| | cDBS+ | aDBS- | | <0.001 | -1.42 | <0.001 | -2.21 |
| | aDBS+ | aDBS- | | <0.001 | -0.83 | <0.001 | 1.92 |
| Sleeping | cDBS+ | aDBS+ | aDBS- | <0.001 | | <0.001 | |
| | cDBS+ | aDBS+ | | <0.001 | 0.98 | <0.001 | -1.36 |
| | cDBS+ | aDBS- | | 0.25 | | <0.001 | -1.85 |
| | aDBS+ | aDBS- | | <0.001 | -0.71 | 0.9 | |
| cDBS+ | Waking | Sleeping | | <0.001 | 1.34 | 0.07 | |
| aDBS+ | Waking | Sleeping | | <0.001 | 1.95 | <0.001 | 1.79 |
| aDBS- | Waking | Sleeping | | <0.001 | 1.92 | 0.25 | |

**Table S1. Statistical comparison in the low beta band.**

Comparison of the daily median and interquartile range of the low-beta amplitude between the three different treatment conditions (cDBS+, aDBS+, and aDBS-) and the two different activity levels (waking and sleeping). The significance level was set to 0.05. Effect size was calculated using Robust Cohen's distance (rCoh, see Star Methods).
Abbreviations: a, adaptive; c, conventional; DBS+, with dopaminergic medication; DBS-, without dopaminergic medication.

| | Factors | | | Median F(2,296)=9.92, p<0.001 | | Interquartile range F(2,592)=29.95, p<0.001 | |
|---|---|---|---|---|---|---|---|
| | **Factor 1** | **Factor 2** | **Factor 3** | **p-value** | **rCoh** | **p-value** | **rCoh** |
| Waking | cDBS+ | aDBS+ | aDBS- | <0.001 | | <0.001 | |
| | cDBS+ | aDBS+ | | <0.001 | 1.91 | <0.001 | -2.49 |
| | cDBS+ | aDBS- | | 0.17 | | 0.0013 | -1.16 |
| | aDBS+ | aDBS- | | <0.001 | -1.36 | <0.001 | 1.40 |
| Sleeping | cDBS+ | aDBS+ | aDBS- | <0.001 | | 0.37 | |
| | cDBS+ | aDBS+ | | <0.001 | 0.80 | | |
| | cDBS+ | aDBS- | | 0.71 | | | |
| | aDBS+ | aDBS- | | 0.003 | -0.54 | | |
| cDBS+ | Waking | Sleeping | | <0.001 | 1.20 | 0.87 | 0.02 |
| aDBS+ | Waking | Sleeping | | <0.001 | 0.74 | <0.001 | 2.22 |
| aDBS- | Waking | Sleeping | | <0.001 | 1.63 | <0.001 | 0.84 |

**Table S2. Statistical comparison in the high beta band.**

Comparison of the daily median and interquartile ranges of the high beta amplitude between the three different treatment conditions (cDBS+, aDBS+, and aDBS-) and the two different activity levels (waking and sleeping). The significance level was set to 0.05. Effect size was calculated using Robust Cohen's distance (rCoh, see Star Methods).
Abbreviations: a, adaptive; c, conventional; DBS+, with dopaminergic medication; DBS-, without dopaminergic medication.
